# Assessing the Impact of the Application of PMSI Data on Enrollment Efficiency, Retention, and Therapeutic Area Performance in Clinical Trials

**DOI:** 10.1101/2025.04.18.25326069

**Authors:** Raimundo Gordejuela, Sneha Keerthi Nama Ravi, Eva Llamas, Ahmad Wali, Aurélie Lampuré, Bernadette Collins, Nathan Sommerford

## Abstract

**Objective:** This study evaluates whether the use of Programme de Médicalisation des Systèmes d’Information (PMSI) data in France positively impacts clinical trial performance or not. Specifically, we compared enrollment performance and dropout rates across therapy areas between PMSI-supported and non-PMSI supported clinical trials.

**Methods:** A retrospective analysis was conducted using enrollment and dropout data for clinical trials conducted between Jan 2019 and Oct 2024, across multiple therapy areas. Comparative statistics and visualizations were used to assess the difference in enrollment performance and dropout rates between PMSI-supported and non-PMSI supported trials.

**Results:** PMSI supported trials demonstrated higher median enrollment rates and fewer outliers across several therapy areas, particularly Infectious Diseases, Gastrointestinal and Dermatology. For Cardiovascular and Oncology trials, the median enrollment rates for PMSI supported trials were slightly higher than the non-PMSI supported trials, indicating a smaller impact on trials in these therapy areas. Specifically, PMSI-supported studies showed enrollment rates that were between 5% and 238% higher than non-PMSI-supported studies, depending on the therapy area. Also, the dropout rates were also lower in PMSI supported trials.

**Conclusion:** The application of PMSI data appears to be associated with improved trial performance in terms of enrollment efficiency, fewer outliers, and consistent trial execution particularly in select therapy areas, namely Infectious Diseases, Gastrointestinal and Dermatology. These findings support the strategic use of PMSI data to enhance trial success and patient recruitment in France.

## Introduction

Successful recruitment is a vital component of clinical trials, as it ensures the validity and reliability of results. Efficient recruitment helps maintain timelines, control costs and ultimately contributes to the success of the trials. Inadequate recruitment of eligible participants or low participants accrual rates for a clinical trial can lead to substantial delays, financial setbacks, diminished validity of the study’s findings or premature termination of clinical trials ^1,2,3,4^.

The clinical trial landscape in France is highly competitive, bolstered by a strong research infrastructure and a robust regulatory framework. Collaboration between public institutions and private companies, along with government support, accelerates trial development. France’s diverse population provides a relevant patient pool from which to recruit for trials, while the adoption of innovative technologies enhances trial efficiency. Major pharmaceutical and biotech firms also play a significant role, contributing to a dynamic environment by contributing to innovative drug development and funding research. However, in recent years France has seen a decline in the number of clinical trials being conducted in the country, due to regulatory challenges, competition from other countries, economic constraints, and the impact of the COVID-19 pandemic ^5,6^.

Despite the decline in the number of trials being conducted in France, the country is still ranked third among European countries for participation in international clinical trials, particularly excelling in oncology, where it ranks second in Europe^7,8^ and ranks fourth for early-stage trials^9.^

France is an ideal environment for conducting clinical trials as it has the second largest population among all countries in the European Union and is a leading country in the field of health with a strong universal healthcare system, global health security, research, and innovation. It has dedicated medical infrastructures and specialized expertise, such as oncology centers, clinical research centers, national clusters, and networks, has experience in designing and running complex innovative trials and data management and has improved speed of enrollment in trials^10^. However, the country faces stiff competition from Spain, Germany, and the United Kingdom across therapeutic areas. Despite efforts to streamline processes, it still takes an average of 160 days to initiate a clinical trial in France, highlighting the need for further improvements. With upcoming European pharmaceutical legislation set to impact the region’s attractiveness for clinical research, the future of France’s position in the global landscape is uncertain.

### Challenges in Ensuring Diversity in Clinical Trials in France

Including a diverse group of participants in clinical trials is crucial for making trial findings applicable to a broader population. However, the collection of ethnicity data is generally forbidden in France, with few exceptions, since the French Constitutional Council prohibits “the processing for the purposes of measuring personal data revealing directly or indirectly the racial or ethnic origin of people, as well as the introduction of variables on race or religion in administrative files.^11^” This restriction limits the ability to assess how treatment works across diverse populations and may reduce the inclusivity and generalizability of real-world data for the French population. As a result, certain patient subgroups may be underrepresented.

### Streamlining Clinical Trials: From CTD to ETR/ Evolution of Clinical Trial Regulations in France and the EU

Until January 2022, clinical trials of medicinal products in the European Union were governed by the Clinical Trials Directive (CTD). In France, French Public Health Code (FPHC) sets the legislative and regulatory framework for all trials involving human subjects. All relevant provisions earlier proposed by the CTD that were applicable to clinical trials were transposed and complemented in The French Public Health Code (FPHC)^12^.

In January 2022, the new European Trial Regulation (ETR) was implemented to promote clinical research in Europe. This new trial regulation is directly applicable in France and governs interventional clinical trials on medicinal products. The regulation harmonizes the processes for assessment and supervision of clinical trials throughout the EU. Prior to the Regulation, clinical trial sponsors had to submit clinical trial applications separately to national competent authorities and ethics committees in each country to gain regulatory approval to run a clinical trial.^12^

The ETR enables sponsors to submit one online application via a single online platform known as the Clinical Trials Information System (CTIS) for approval to run a clinical trial in several European countries, making it more efficient to carry out such multi-national trials. The ETR also makes it more efficient for EU Member States to evaluate and authorize such applications together, via the Clinical Trials Information System. The purpose is to foster innovation and research in the EU, facilitating the conduct of larger clinical trials in multiple EU Member States/EEA countries^13^.

This new regulation introduces several key changes. It simplifies the Clinical Trials (CT) application process by eliminating the need for separate submissions to each national authority. It improves the approval turnaround time (TAT), granting quicker access to innovative treatments and therapies to patients. It mandates public access to clinical trial data through CTIS, building trust and encouraging patient participation in France. It simplifies procedures and promotes a cohesive regulatory environment. This makes France more appealing to sponsors, potentially increasing the demand for French clinical research and providing patients quicker access to innovative treatments ^12,13^.

Additionally, the Health innovation 2030 Plan, announced by the French President in June 2021 aims to increase the number of clinical trials and enrolled patients by fast-tracking authorizations, mobilizing additional financial resources and improving the call for projects^14^.

These recent regulation updates and new innovations could help to stop the decline in trials being conducted in France.

### PMSI Database

Programme de Médicalisation des Systèmes d’Information (PMSI) is a French national hospital discharge database that covers data on all hospital stays in France, both in public and private settings. The database is processed and managed by the Technical Agency for Hospitalization Information (ATIH) and contains medical, administrative and patient information^15,16^.

The PMSI was created to improve the management and financing of hospitals in France. It was designed to collect detailed information on hospital activities, including patient demographics, diagnoses, medical procedures, consumption of certain medications dispensed in a hospital setting and hospital costs. PMSI contains data on all hospitalizations in medicine, chirurgie (surgery) and obstetrics (MCO), in follow-up and rehabilitation care (SSR), in psychiatry (RIM-P) and home hospitalizations (HAD)^16^.

PMSI is part of the larger French National Health Insurance Information System, known as Système National d’Information Inter-Régime de l’Assurance Maladie (SNIIRAM). SNIIRAM includes data from various sources, such as hospital discharge records, health insurance claims, demographic information, and death records^17^.

The data collected through PMSI can be used for various purposes, such as to analyze and improve hospital management, optimize hospital finance, enhance quality of care, shape health policies, allocate resources and support clinical research.

PMSI is a valuable resource for clinical research, allowing long term follow-up of patients that helps to support clinical trial recruitment, study patient care pathways and treatment outcomes.

In clinical research, the performance metrics of studies such as Enrollment Rate (ER) and Dropout Rate (DR) are critical indicators of a study’s efficiency and effectiveness. These metrics not only impact the timeline and cost of clinical trials but also influence the quality and reliability of the study outcomes. The study outlined in this paper focused on analyzing the differences in these performance metrics for studies where data from the Programme de Médicalisation des Systèmes d’Information (PMSI) in France was used versus studies where the data was not used.

IQVIA leverages access to extensive datasets, including the French National Health Data System/ Système National des Données de Santé (SNDS), PMSI, IQVIA Retail, Hospital, and Longitudinal Prescription (LRx) data, which collectively cover healthcare details for 66 million people in France^18^. These resources enable IQVIA to support clinical trial development through real-world evidence (RWE), enhancing patient outcomes and informed healthcare decisions. By employing innovative solutions such as decentralized trials, AI-driven analytics, and predictive biomarkers, IQVIA optimizes patient recruitment and retention, reduces burdens on participants and sites, and boosts trial efficiency. Its comprehensive services, from study design to data management, accelerates decision-making and mitigates risks and ultimately advances clinical research in France, by expediting the delivery of transformative therapies^19^.

Our goal was to analyze and determine whether there is a statistically significant difference in the performance of PMSI-supported clinical trials compared to non-PMSI-supported clinical trials. Specifically, we aimed to investigate:

- The Enrollment Rates for PMSI-supported and non-PMSI-supported studies.
- The Dropout Rates for PMSI-supported and non-PMSI-supported studies.

The dataset considered for analysis includes information on Enrollment Rate (ER) and Dropout Rate (DR) from IQVIA Past Performance Report (PPR) among others (Citeline and Clinical Trials.gov), for studies both supported and not supported by PMSI. The Past Performance Report (PPR) is an IQVIA Dashboard summarizing data and analysis collected from our internal CTMS (Clinical Trial Management System).

## Methods

### Data Preparation

Data spanning Jan 2019 to Oct 2024 was imported from IQVIA internal platforms (i.e., Strategy Workbench and Past Performance Review), along with internal France PMSI logs. The data was then cleaned and processed for analysis. For example, numerical columns were converted to their appropriate data types to ensure consistency and ease of use. To reduce the impact of extreme values and minimize bias in statistical estimates, outliers were identified and removed using the simple and robust interquartile range (IQR) method with a threshold of +-1.5 that relies on the median and quartiles. Finally, a subset of the top therapeutic areas was selected on which to conduct the statistical analysis. These therapeutic areas were selected based on having had a high volume of trials conducted by IQVIA France, to ensure the data used was robust and that meaningful insights could be derived.

### Exploratory Data Analysis (EDA)

An initial exploratory data analysis (EDA) was conducted to examine the distribution of the target variables and their relationship with the PMSI variables. Descriptive statistics and visualizations, including histograms, box plots and violin plots were used to compare the distributions and central tendencies of key metrics between PMSI supported and non-PMSI supported studies. These analyses provided insights into potential patterns within the dataset.

### Statistical Analysis

Several statistical methods were employed to test the hypothesis that the IQVIA France enrollment from PPR for PMSI supported studies differs significantly from non-PMSI supported studies. Given the non-normal distribution of data, the Mann-Whitney U test, a non-parametric statistical test was considered to compare the two groups.

## Results

### Data Preparation and Exploratory Data Analysis (EDA)

Initial exploratory data analysis revealed notable differences in the distribution of Enrollment Rate as per IQVIA Past Performance Review and Dropout Rate between PMSI supported and non-PMSI supported studies.

Descriptive statistics showed that PMSI supported studies had higher mean and median IQVIA Enrollment Rate Past Performance Review (ER PPR) values compared to non-PMSI supported studies. Additionally, specific therapeutic areas, including Gastrointestinal, Dermatology, Infectious Diseases, Oncology, and Cardiovascular, demonstrated consistently higher IQVIA ER PPR values in PMSI supported studies.

### IQVIA Enrollment Rate as per Past Performance Review data for PMSI and non-PMSI supported. Interquartile Range (IQR) Analysis

The IQR for ER PPR was found to be lower in PMSI supported studies compared to non-PMSI supported studies. A lower IQR indicates that the middle 50% of data points are closer to each other, suggesting lower variability and greater consistency in enrollment rates for PMSI supported studies.

**Plot 1.**
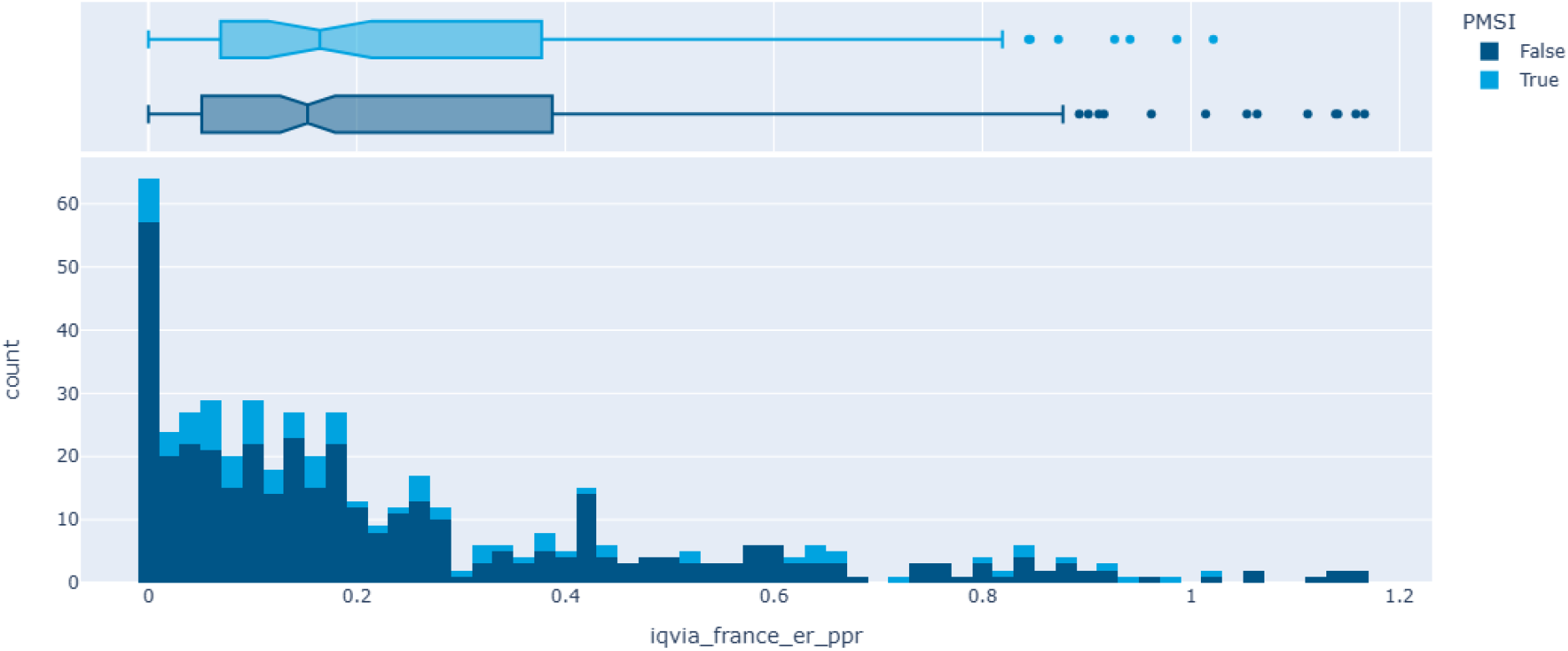
Comparison of enrollment rate performance for PMSI vs non-PMSI supported trials.

The boxplot and histogram illustrate the distribution and frequency of enrollment rates for all IQVIA PMSI supported and IQVIA non-PMSI supported trials.

PMSI supported trials have slightly higher median enrollment rates with a more compact distribution, indicating less internal variability and consistent performance. These trials also show more high-end outliers, reflecting stronger enrollment or trials with exceptional performance.

In contrast, non-PMSI supported trials display a lower median and greater internal variability, with more trials concentrated at a low enrollment rate range of 0-0.1.

Consistency in performance metrics is crucial for clinical trials as it ensures predictability and reliability in study outcomes. Lower variability in enrollment rates can contribute to more stable and reproducible results, which is essential for validating clinical findings.

**Table 1** presents the mean and median ER PPR values for PMSI supported and non-PMSI supported studies across different therapeutic areas.

**Table 1:** IQVIA ER PPR statistics for PMSI supported and non-PMSI supported studies.

| Therapeutic Area | PMSI supported studies ER (Mean) | PMSI supported studies ER (Median) | non-PMSI supported studies ER (Mean) | non-PMSI supported studies ER (Median) | Median ER % increase in PMSI supported studies |
| --- | --- | --- | --- | --- | --- |
| Oncology | 0,2435 | 0,1817 | 0,2475 | 0,1731 | 5% |
| Cardiovascular | 0,2783 | 0,1522 | 0,1622 | 0,0704 | 116% |
| Gastrointestinal | 0,337 | 0,2771 | 0,2075 | 0,1217 | 128% |
| Dermatology | 0,5159 | 0,5159 | 0,2427 | 0,1527 | 238% |
| Infectious Diseases | 0,5313 | 0,5281 | 0,1898 | 0,1682 | 214% |

The data suggests that PMSI supported studies exhibit a higher and more stable enrollment rate compared to non-PMSI supported studies across multiple therapeutic areas. Specifically, PMSI supported studies demonstrated enrollment rates that were between 5% and 238% higher than the non-PMSI supported studies, depending on the therapy area.

**Plot 2.**
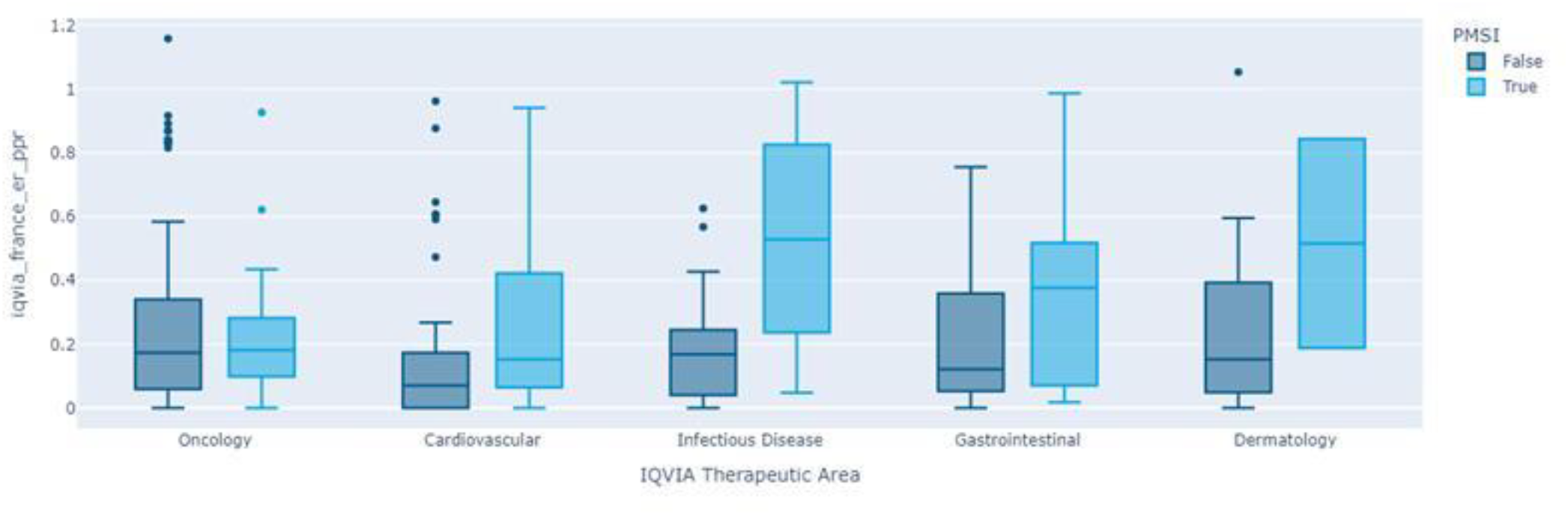
Enrollment rate distributions across top five therapy areas.

The boxplot compares enrollment rates across five therapy areas, stratified by PMSI support status.

PMSI supported trials showed a higher median above 0.5, compared to non-supported PMSI trials which had a median of less than 0.2. The PSMI group also exhibited a wider range of enrollment extending up to 1.0, indicating consistently strong enrollment performance.

For Dermatology, PMSI supported trials have a notably higher median range, approximately 0.45-0.5, while non-PMSI supported trials had a median enrollment of less than 0.2. Also, the upper whisker for PMSI supported trials extends up to 0.9, indicating strong enrollment performance.

For Gastrointestinal, the median enrollment for PMSI supported trials was a little lower than 0.4, but clearly higher than the non-PMSI supported trials.

For Cardiovascular and Oncology trials, the median for PMSI supported trials is slightly higher than for non-PMSI supported trials, suggesting a relatively smaller impact of PMSI support for trials in these therapy areas.

Across therapy areas, PMSI support appears to be associated with consistent or improved enrollment performance.

### Statistical Analysis ER PPR

A Mann-Whitney U test was conducted to compare IQVIA ER PPR values between PMSI supported and non-PMSI supported studies. The results are summarized in Table 2.

**Table 2:** Mann-Whitney U test results for IQVIA ER PPR.

| Statistic | Value |
| --- | --- |
| U-Statistic | 4667.0 |
| p-value | 0.028 |

The Mann-Whitney U test with alpha level of 0.05 confirmed a statistically significant difference in ER PPR between PMSI supported and non-PMSI supported studies, with PMSI supported studies demonstrating higher enrollment performance.

### Exploratory Analysis of Dropout Rate

Exploratory Data Analysis of dropout rates also indicated a difference between PMSI supported and non-PMSI supported studies.

**Table 3:**
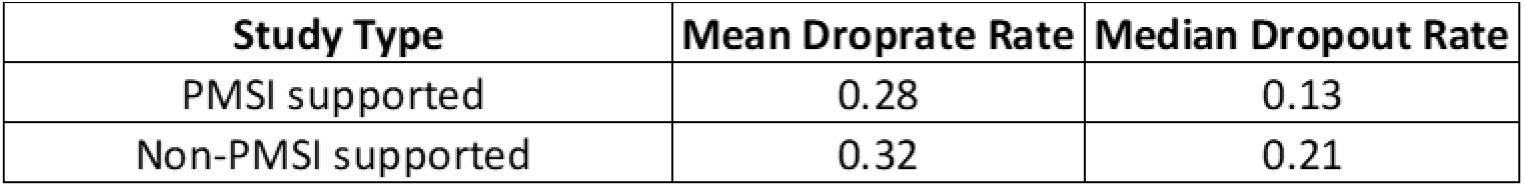
Dropout Rate Statistics for PMSI supported and non-PMSI supported studies.

| Study Type | Mean Droprate Rate | Median Dropout Rate |
| --- | --- | --- |
| PMSI supported | 0.28 | 0.13 |
| Non-PMSI supported | 0.32 | 0.21 |

The results suggest that PMSI supported studies have lower dropout rates compared to non-PMSI supported studies. Lower dropout rates are essential for maintaining the integrity of study data, as they help reduce bias that may arise from incomplete data. The findings imply that PMSI supported studies may benefit from better participant retention strategies, ultimately contributing to the success and reliability of clinical trials.

## Discussion

In this study, we analyzed the performance metrics of clinical trials supported by the PMSI compared to those without PMSI support.

The data indicates relatively greater impact of PMSI supported trails in therapy areas such as Gastrointestinal, Dermatology, Infectious Diseases, Cardiology and Oncology, this may be influenced by several interrelated factors. Patient populations in these therapy areas may follow more defined care pathways, with earlier and more consistent interaction with healthcare providers, which can support timely identification and recruitment into the clinical trials. The study designs and protocols in these therapeutic domains could be less complex, enhancing feasibility for participants. Prior success and established experience with PMSI supported studies in these therapy areas may foster continued confidence and collaboration, contributing to enhanced trial performance observed. Hence, the structured and comprehensive data provided by PMSI plays a crucial role in enhancing participant recruitment and retention in clinical trials.

One of the notable findings in this study is the lower variability in enrollment rates for PMSI-supported studies. This consistency is essential for clinical trials as it ensures predictability and reliability in study outcomes. The reduced variability can be attributed to the robust data management and patient tracking capabilities of PMSI, which streamline the recruitment process and minimize delays. Additionally, the lower dropout rates observed in PMSI-supported trials highlight the effectiveness of PMSI in maintaining participant engagement and reducing attrition, thereby preserving the integrity of the study data.

The implementation of the new European Trial Regulation and the Health Innovation 2030 Plan in France further underscores the importance of leveraging advanced data systems like PMSI. These initiatives aim to streamline clinical trial processes, enhance regulatory efficiency, and mobilize additional resources to support clinical research. By integrating PMSI data with these regulatory frameworks, France can potentially reverse the declining trend in clinical trials and strengthen its position as a leading hub for clinical research in Europe.

## Conclusion

In conclusion, PMSI-supported trials consistently demonstrate enhanced enrollment performance compared to non-PMSI supported trials, across various therapy areas, and with overall medians across the top five therapy areas ranging from 5% to 238%. For Dermatology, Infectious Diseases and Gastrointestinal studies, PMSI trials showed significantly higher median enrollment rates, with Dermatology reaching up to 0.9 pat/site/month. Cardiovascular and Oncology trials also benefited, though to a lesser extent. This consistent improvement emphasizes the value of PMSI data in optimizing trial efficiency and reducing dropout rates, ultimately accelerating the development of innovative therapies. By leveraging PMSI’s robust datasets, clinical research gains both reliability and speed, reinforcing the critical role of data-driven strategies in transforming healthcare and delivering life-changing treatments to patients.

Further research should continue exploring these relationships and identifying strategies to optimize study performance across all therapeutic areas, ensuring that clinical trials remain both effective and patient centered.

## Limitations

This study has limitations that should be considered when interpreting the findings:

### 1. Lack of country level enrollment data from clinical trial databases

Clinical trial databases such as Citeline and clinical trials.gov do not report enrollment rates at a country level. This limitation made it challenging to conduct direct comparisons between PMSI-supported and non-PMSI supported studies at overall clinical trial level for France. As a result, our analysis is constrained to IQVIA project level insights rather than detailed country specific enrollment trends.

### 2. Enrollment Rate Data Constraints

The enrollment rate data reported in Citeline and clinical trials.gov is available as aggregated enrollment rates across countries for each trial, rather than at a country level. To analyze country specific enrollment rates, we relied exclusively on internal resources and proprietary data that collects clinical trial data at a country level for each trial, which limits the sample size and hence may not represent global trends.

### 3. Insufficient data to test a hypothesis at indication level

The data was sufficient to test the hypothesis at a therapeutic area level but restricted our ability to test the hypothesis at a specific indication level.

### 4. Variability in Study Design

Differences in study designs, sample sizes and therapeutic areas may contribute to variability in enrollment and dropout rates. The data was treated, but unmeasured confounders could influence the observed difference between PMSI supported and non-PMSI supported studies.

### 5. Potential Confounding Factors

The observed advantages in recruitment and retention in PMSI supported trials may be influenced by confounding factors such as differences in trial complexity, site capabilities, and patient pathways. Additionally, variations in data quality and coding practices across therapy areas could impact patient identification and follow-up. These factors may limit direct comparisons with non-PMSI supported trials.

## Future Work

### 1. Investigate additional performance metrics

Beyond enrollment and dropout rates, future studies should examine other performance indicators like screen failure rates, enrollment duration and the total number of subjects enrolled. Analyzing these metrics will provide a more comprehensive understanding of how PMSI support influences clinical trial efficiency.

### 2. Longitudinal Analysis

Conducting a longitudinal analysis will help identify and depict trends over time and assess whether the effects of PMSI support on study performance remain consistent across different time periods. This approach can generate insights to support long term improvements and help in conducting informed future trial planning.

## Data Availability

All data produced in the present study are available upon reasonable request to the authors

## Abbreviations

ATIH: Technical Agency for Hospitalization Information
CT: Clinical Trials
CTD: Clinical Trials Directive
CTIS: Clinical Trials Information System
DR: Dropout Rate
EDA: Exploratory Data Analysis
ER: Enrollment Rate
ETR: European Trial Regulation
EU: European Union
FPHC: French Public Health Code
IQR: Interquartile Range
LRx: Longitudinal Prescription data
MCO: Medicine, Chirurgie (Surgery) and Obstetrics
PMSI: Programme de Médicalisation des Systèmes d’Information
PPR: Past Performance Review
RWE: Real-World Evidence
SNDS: Système National des Données de Santé/French National Health Data System
SNIIRAM: Système National d’Information Inter-Régime de l’Assurance Maladie
TAT: Turnaround Time

